# Triage tool for suspected COVID-19 patients in the emergency room: AIFELL score

**DOI:** 10.1101/2020.05.09.20096834

**Authors:** Ian Levenfus, Enrico Ullmann, Edouard Battegay, Macé M. Schuurmans

## Abstract

Clinical prediction scores support the assessment of patients in the emergency setting to determine the need for further diagnostic and therapeutic steps. During the current COVID-19 pandemic, physicians in emergency rooms (ER) of many hospitals have a considerably higher patient load and need to decide within a short time frame whom to hospitalize. Based on our clinical experiences in dealing with COVID-19 patients at the University Hospital Zurich, we created a triage score with the acronym AIFELL consisting of clinical, radiological and laboratory findings.

The score was then evaluated in a retrospective analysis of 122 consecutive patients with suspected COVID-19 from March until mid-April 2020. Descriptive statistics, Student’s t-test, ANOVA and Scheffe’s post hoc analysis confirmed the diagnostic power of the score. The results suggest that the AIFELL score has potential as a triage tool in the ER setting intended to select probable COVID-19 cases for hospitalization in spontaneously presenting or referred patients with acute respiratory symptoms.

## Introduction

Due to the worldwide spread of SARS-CoV-2 and rapidly increasing numbers of infections, the novel coronavirus became a considerable strain for emergency rooms (ER), especially when several suspected cases with unspecific general or respiratory symptoms arrive at the same time. Identification of more critical patients in the ER for hospitalization is a challenge since the detection of SARS-CoV-2 in nasopharyngeal swabs by quantitative polymerase chain reaction (qPCR) still requires many hours (> 6 hours in our setting). Therefore, the qPCR result currently cannot be used in the frontline setting to decide whom to hospitalize and who can be managed as an outpatient. Rapid point of care tests for SARS-CoV-2 are being developed, but are currently not validated for routine use.[1]

As a frontline physician, whose task was to evaluate and triage patients arriving with symptoms suggesting COVID-19 in the ER coronavirus unit of the University Hospital Zurich, the first author was confronted with the problem of whom to choose for hospitalization due to probable COVID-19 and whom to discharge whilst the qPCR results of the swab were pending.

The hospitalization criterion was to select patients at risk for developing more severe symptoms leading to respiratory failure (COVID Stages II or III [2]). During clinical routine work in the frontline unit, the question of a score arose to support the triage process and assist other physicians in similar situations.

## Methods

We therefore followed up consecutively hospitalized patients with proven COVID-19 in order to determine initial features which may help distinguishing probable COVID-19 cases from other respiratory problems. From 30 personally encountered consecutively hospitalized patients with proven COVID-19 studied initially and evidence obtained from literature searches using the keywords “COVID-19”, “SARS-CoV-2”, “laboratory” and “patients” in PubMed[3–6], the relevant components and cut-off values became more evident and a simple score was created based on typical clinical information available in our ER.

The AIFELL score includes an **<u>A</u>**ltered sense of smell/taste, **I**nflammation (C-reactive protein ≥ 30 mg/l), radiological **<u>I</u>**nfiltrates, **<u>F</u>**ever (≥ 38.0°C), **<u>E</u>**levated **<u>L</u>**actate dehydrogenase (LDH) levels (> 400 U/l) and **<u>L</u>**ymphocytopenia (absolute count < 1.45 G/l). The score is calculated by counting the number of criteria met at initial presentation in the ER, whereas each criterion equals one point (Score range 0 to 6 points).

To assess the score, we applied it retrospectively to consecutive patients with suspected COVID-19 admitted via the ER from March until mid-April 2020. Only those cases evaluated with chest imaging and a blood test including at least two of the three considered blood parameters at presentation in the ER, who did not decline the general research consent, were included. Of 122 patients with suspected COVID-19, 52 cases turned out to have other respiratory problems (Table 1). SARS-CoV-2 positive patients (N=70) were classified according to the stages suggested by Siddiqi and Mehra (Figure 1)[2]. The study was approved by the Institutional Review Board (Ethics Committee of the Canton of Zurich, Nr. 2020-00854).

**Table 1:**
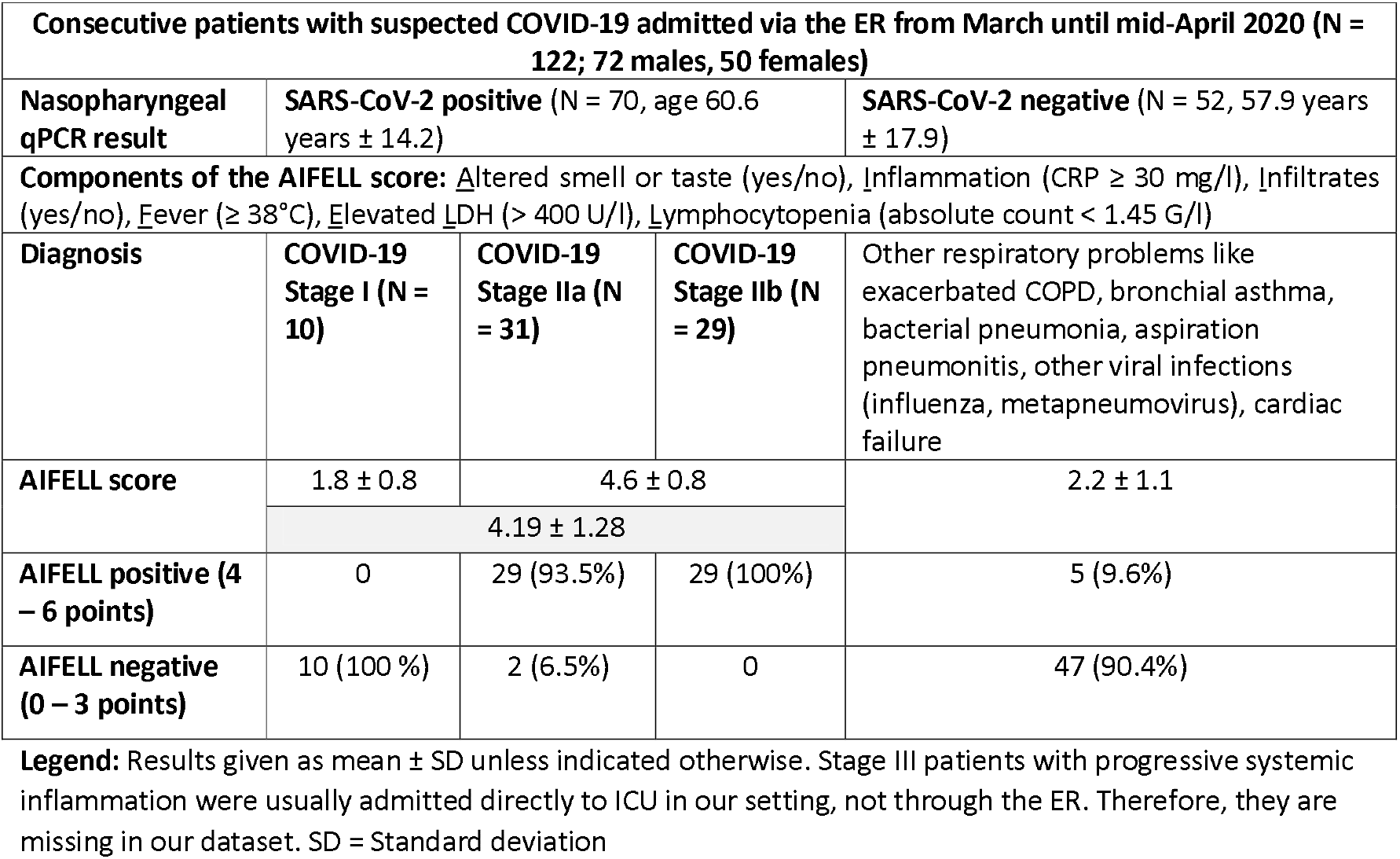
Distribution of enclosed patients and their clinically assigned AIFELL scores. Legend: Results given as mean ± SD unless indicated otherwise. Stage III patients with progressive systemic inflammation were usually admitted directly to ICU in our setting, not through the ER. Therefore, they are missing in our dataset. SD = Standard deviation

**Figure 1:**
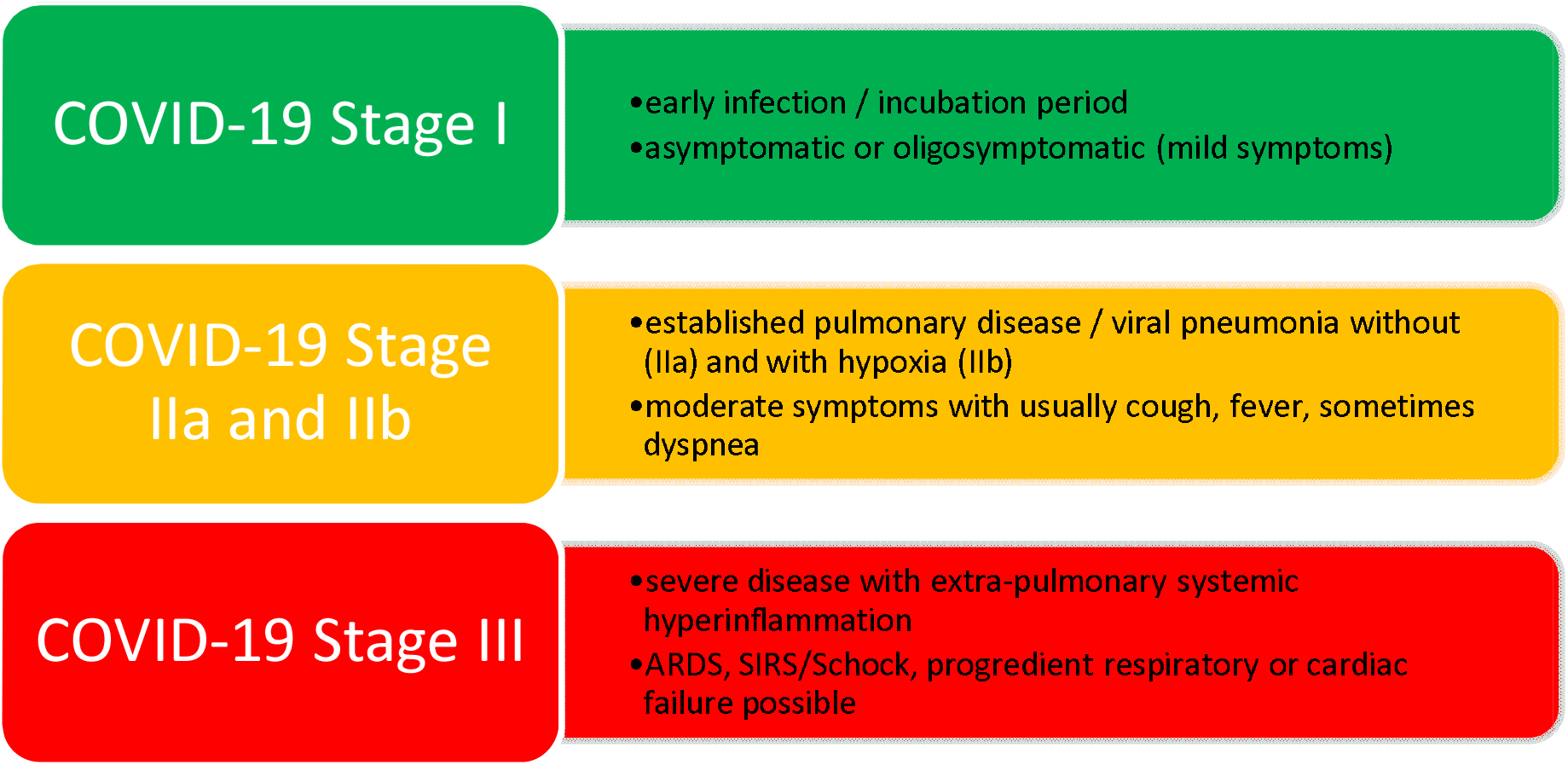
COVID-19 Stages as suggested by Siddiqi und Mehra.[2] In COVID-19 Stage hospitalization should be considered. Stage III patients usually need ICU admission.

After testing for normal distribution and standardized outliers, we created a new variable named “paraclinical measurements” by including the z-standardized mean values of LDH, C-reactive protein (CRP), inversely poled serum lymphocytes and auricular body temperature. We afterwards summarized this variable with lung infiltrates seen by imaging and alterations of smell/taste indicated by patient history to get our predictor score named “AIFELL”. Student’s t-tests and an ANOVA with Scheffe’s post hoc tests were performed for group comparisons. For all analyses, we used MS Excel 2016 and SPSS 24.

## Results

The mean age of our SARS-CoV2 positive subjects (N=70) was 60.6 years ± 14.2 (Standard deviation) vs. 57.9 years ± 17.9 of our SARS-CoV2 negative subjects (N=52; t=.908; p=.37). There were significantly different AIFELL scores (t=5.77, p<0.001) between SARS-CoV-2 positive patients (mean=2.43 ± 0.15 Standard error) and SARS-CoV-2 negative patients (mean=1.30 ± 0.12) (Figure 2).

**Figure 2:**
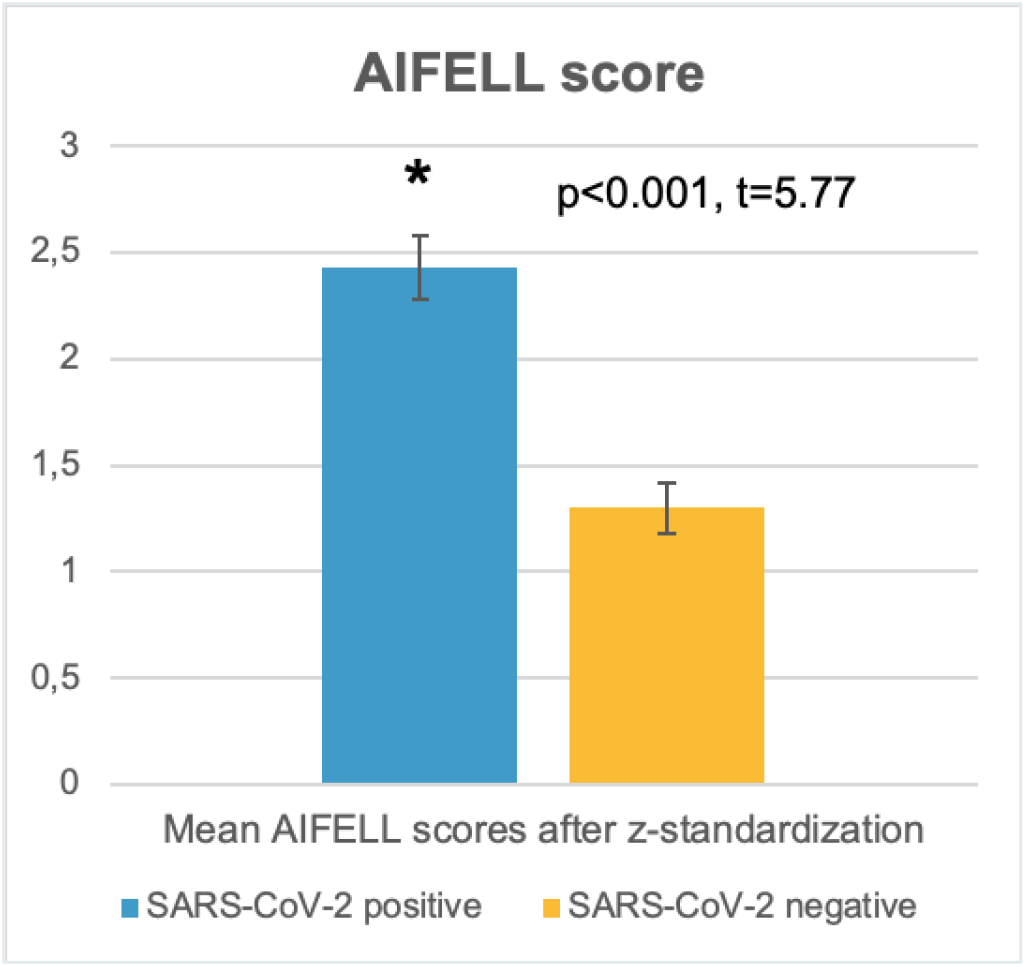
Mean AIFELL scores after z-standardization in SARS-CoV-2 positive (2.43 ± 0.15 Standard error) and SARS-CoV2 negative patients (1.30 ± 0.12)

A score of ≥ 4 points/criteria met at presentation was highly associated with qPCR-based SARS-CoV-2 detection in nasopharyngeal swabs and presence of symptomatic COVID-19 (Stages II or III), thus justifying hospitalization. Scores between 0 and 3 were associated with other respiratory conditions (Table 1).

The ANOVA and post hoc tests verified significant differences of evaluated score component values between different score counts (Table 2).

**Table 2:**
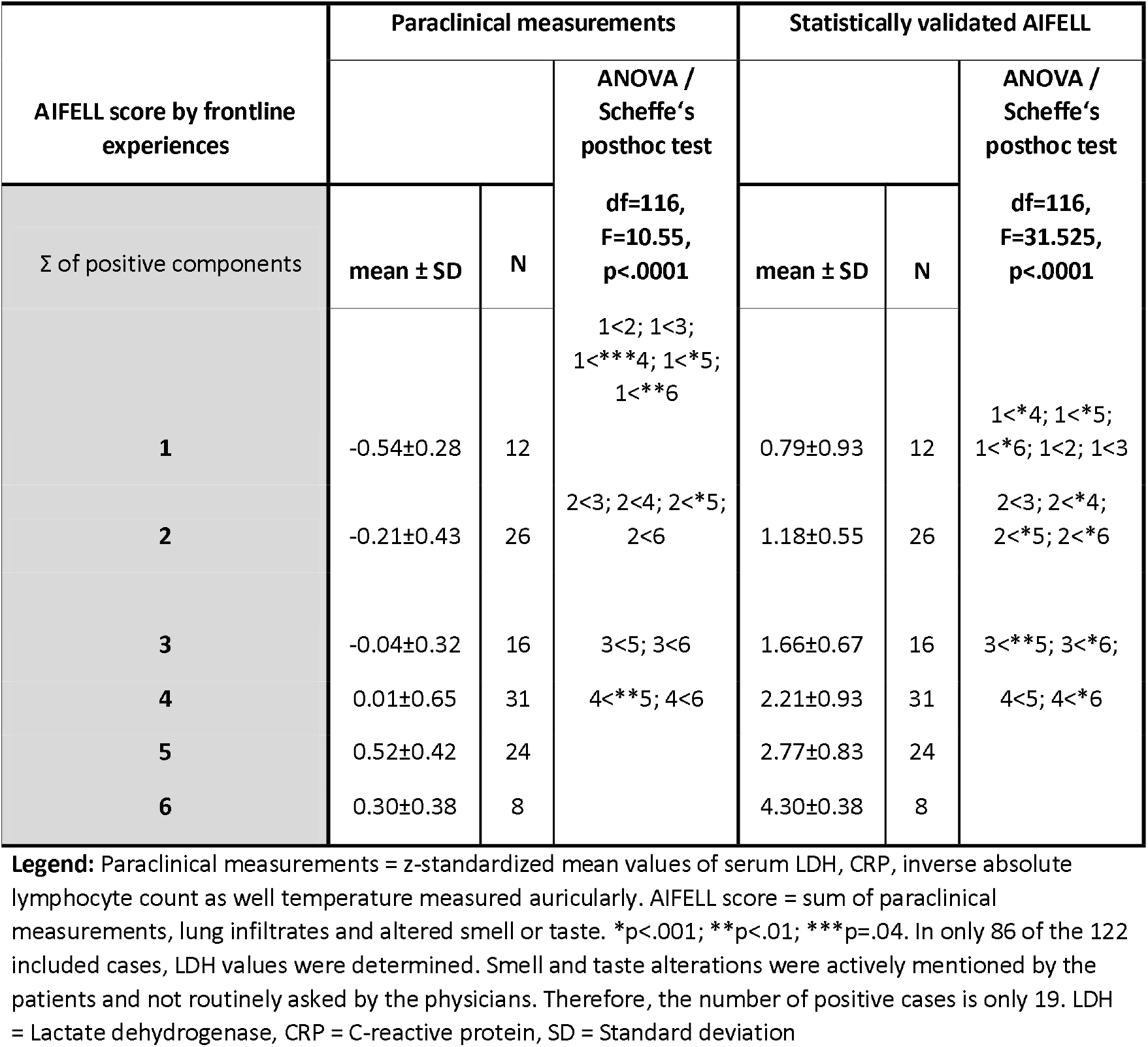
Group differences between AIFELL created by clinical information and statistically verified AIFELL. Legend: Paraclinical measurements = z-standardized mean values of serum LDH, CRP, inverse absolute lymphocyte count as well temperature measured auricularly. AIFELL score = sum of paraclinical measurements, lung infiltrates and altered smell or taste. *p<.001; **p<.01; ***p=.04. In only 86 of the 122 included cases, LDH values were determined. Smell and taste alterations were actively mentioned by the patients and not routinely asked by the physicians. Therefore, the number of positive cases is only 19. LDH = Lactate dehydrogenase, CRP = C-reactive protein, SD = Standard deviation

## Discussion

Based on the evaluation of the initial data of 30 patients, we generated the AIFELL score as a simple triage instrument for the ER setting consisting of frequently available elements like patient symptoms (fever, altered smell and taste), laboratory values (differential blood count, CRP, LDH) and imaging. Afterwards, we evaluated its diagnostic performance in a larger number of consecutive patients.

A host risk score dealing with comorbidities of COVID-19 patients has been previously published in a Chinese study.[7] Scores regarding hyperinflammation in COVID-19 have also been discussed.[8] However, no ER triage score to identify probable COVID-19 cases in more critical stages (II and III) has been proposed yet. The AIFELL score uses only frequently obtained data usually available both, in the ER and the general practice setting. Other, more sophisticated laboratory parameters such as interleukin (IL)-6[9], soluble IL-2 receptor[10], Ferritin and D-dimers[9] may also be of interest, but are not widely available or more time consuming, thus not practicable in this setting.

During the COVID-19 pandemic and partly scarce medical resources, the score may be useful for selecting symptomatic COVID-19 cases from rather unspecific general or respiratory symptoms in the ER who should immediately get a SARS-CoV-2 swab due to higher probability of the disease. The score is not intended to identify asymptomatic or oligosymptomatic SARS-CoV-2 infections (COVID-19 Stage I).

The major limitation of this work is the single-center evaluation of only a limited number of patients. Due to its retrospective nature, in 36 of all included cases, LDH values were missing as it was not measured routinely on admission. Smell and taste alterations were actively mentioned by the patients and not routinely asked by physicians yet. The strengths are its simplicity, immediate availability as well as wide applicability due to simple components.

The AIFELL score obviously needs to be prospectively applied in larger cohorts of patients to gain more reliable data regarding its diagnostic yield.

## Data Availability

The original raw data is password-protected and stored in protected folders on servers of the University Hospital Zurich and is only accessible by the study team as approved by the IRB. Anonymized data for statistical analysis is also on servers of the University Hospital Zurich.

## Notes

### Competing Interest Statement

The authors have declared no competing interest.

### Funding Statement

No funding was received.

